# SARS-CoV-2 infection in health care workers: a retrospective analysis and model study

**DOI:** 10.1101/2020.03.29.20047159

**Authors:** Yansen Bai, Xuan Wang, Qimin Huang, Han Wang, David Gurarie, Martial Ndeffo-Mbah, Fei Fan, Peng Fu, Mary Ann Horn, Shuai Xu, Anirban Mondal, Xiaobing Jiang, Hongyang Zhao

**Author notes:** **Correspondence:** Xiaobing Jiang, MD, PhD, Department of Neurosurgery, Union Hospital, Tongji Medical College, Huazhong University of Science and Technology, Wuhan 430022, China., Hongyang Zhao, MD, PhD, Department of Neurosurgery, Union Hospital, Tongji Medical College, Huazhong University of Science and Technology, Wuhan 430022, China. These authors contributed equally to this study (Yansen Bai, Xuan Wang, Qimin Huang).

## Abstract

**Background:** There had been a preliminary occurrence of human-to-human transmissions between healthcare workers (HCWs), but risk factors in the susceptibility for COVID-19, and infection patterns among HCWs have largely remained unknown.

**Methods:** Retrospective data collection on demographics, lifestyles, contact status with infected subjects for 118 HCWs (include 12 COVID-19 HCWs) from a single-center. Sleep quality and working pressure were evaluated by Pittsburgh Sleep Quality Index (PSQI) and The Nurse Stress Index (NSI), respectively. Follow-up duration was from Dec 25, 2019, to Feb 15, 2020. Risk factors and transmission models of COVID-19 among HCWs were analyzed and constructed.

**Findings:** A high proportion of COVID-19 HCWs had engaged in night shift-work (75.0% vs. 40.6%) and felt they were working under pressure (66.7% vs. 32.1%) than uninfected HCWs. COVID-19 HCWs had higher total scores of PSQI and NSI than uninfected HCWs. Furthermore, these scores were both positively associated with COVID-19 risk. An individual-based model (IBM) estimated the outbreak duration among HCWs in a non-typical COVID-19 ward at 62-80 days and the basic reproduction number *R*_0_ =1.27 [1.06, 1.61]. By reducing the average contact rate per HCW by a 1.35 factor and susceptibility by a 1.40 factor, we can avoid an outbreak of the basic case among HCWs.

**Interpretation:** Poor sleep quality and high working pressure were positively associated with high risks of COVID-19. A novel IBM of COVID-19 transmission is suitable for simulating different outbreak patterns in a hospital setting.

**Funding:** Fundamental Research Funds for the Central Universities

## Introduction

In December 2019, pneumonia with previously unknown etiology began to spread in Wuhan, Hubei province in China. The causative virus of this disease was soon identified as a novel coronavirus, and it was preliminarily named as 2019 novel coronavirus (2019-nCoV). This virus was later renamed as SARS-CoV-2, and pneumonia it causes was named 2019 novel coronavirus diseases (COVID-19) by World Health Organization (WHO). As with other infectious disease outbreaks, healthcare workers (HCWs) have been at the front line of the fight against COVID-19. However, hospitals are vulnerable to infectious disease spread through rapid patients-HCWs and HCWs-HCWs cross-infection, especially when dealing with a disease of unknown or not well-known etiology as it was the case during the early phase of the COVID-19 outbreak ^1-3^.

At first, the number of new COVID-19 cases in Wuhan city was increasing by more than 5,000 each day. Such pressure resulted in overloading for the health-care systems and increasing rate of contact infection for HCWs. More importantly, the shortage of personal protective equipment (PPE) exacerbates this situation and expands panic. A recent study from the Chinese Center for Disease Control and Prevention showed that a total of 1,716 HCWs had been diagnosed with COVID-19, including 5 deaths by Feb 11, 2020, with a crude case fatality rate of 0.3% ^4^. Additionally, shortage of HCWs resulted in collapse of medical system, even though Wuhan had the high-quality top-notch medical resource. Subsequently, thousands of HCWs from various provinces in China participated in the support of Wuhan epidemic and greatly eased the strain. At present, a serious infection of HCWs has also occurred in the worldwide. Reports from Italy indicated that 20% of responding HCWs were infected with COVID-19.^5^ In Spain, HCWs infected with COVID-19 accounts for around 12% of all confirmed cases.^6^ Therefore, the establishment of protection guideline for HCWs is an important step to fight against COVID-19, and is the most important bridge that prevents the collapse of the medical system and reduces social panic. However, the specific reasons for the infection of HCWs and the failure of protection still need to be further investigated.^7^

To the best of our knowledge, the risk factors in the susceptibility for COVID-19 among HCWs have largely remained unknown, given that there is no existing peer-reviewed literature quantifying the transmissibility of SARS-CoV-2 among HCWs before the date of the outbreak of COVID-19.

In addition, the dynamics of COVID-19 spread among HCWs largely remained unknown. Mathematical modeling of disease transmission allows one to quantify and explain observed infection patterns and trends. Furthermore, such models fitted to data (calibrated and validated) can be used for prediction/control analysis, to explore efficient interventions and design control strategies. Traditional approaches in epidemiological modeling use compartmental models (SEIR), which assume a uniform host population and simple mixing patterns with steady contact rates.^8^ But such models are too crude to account for specifics of COVID-19 in a relatively small host pool. The latter is characterized by highly heterogeneous host populations, disease progress histories, and behavioral (contact) patterns. Our individual-based model (IBM) features SEIR-type infection states, random disease progression and mixing contacts; all are important in a local (nosocomial) transmission setting. One of the important parameters in mathematical models is the basic reproduction number (*R*_0_), which is defined as the average number of secondary infections that arise from a typical primary case in a completely susceptible population. It is useful as a marker for the risk of an outbreak (speed and intensity), and it could be used to assess control effort.^8,9^

In the present study, a retrospective study of 118 HCWs, including 12 confirmed COVID-19 HCWs from a single-center of case-control series was conducted. Their information before the phase of the outbreak of COVID-19, including epidemiological, demographical, lifestyles were collected. We aim to investigate the risk factors that play roles in the susceptibility of HCWs to COVID-19. Further, we designed a novel individual-based model (IBM) to simulate the dynamics of COVID-19 spread through person-to-person contacts among HCWs, and explored its implications for assessment of outcomes and control implications.

## Materials and methods

### Study Design and Participants

We carried out this single-center study in the Department of Neurosurgery, Union Hospital, Tongji Medical College, Huazhong University of Science and Technology, Wuhan, China. 14 of 171 HCWs (an infection rate of 8.19%) in this single-center were infected with SARS-CoV-2 by a hospitalized patient who was later diagnosed with COVID-19 and defined as the index case. 12 of the 14 COVID-19 HCWs with complete questionnaire data were enrolled in this study. The overview of the transmission of SARS-CoV-2 from this index case to 12 HCWs was shown in Figure 1. This study was approved by the institutional ethics board of Union Hospital, Tongji Medical College, Huazhong University of Science and Technology (No. 20200029).

**Figure 1.**
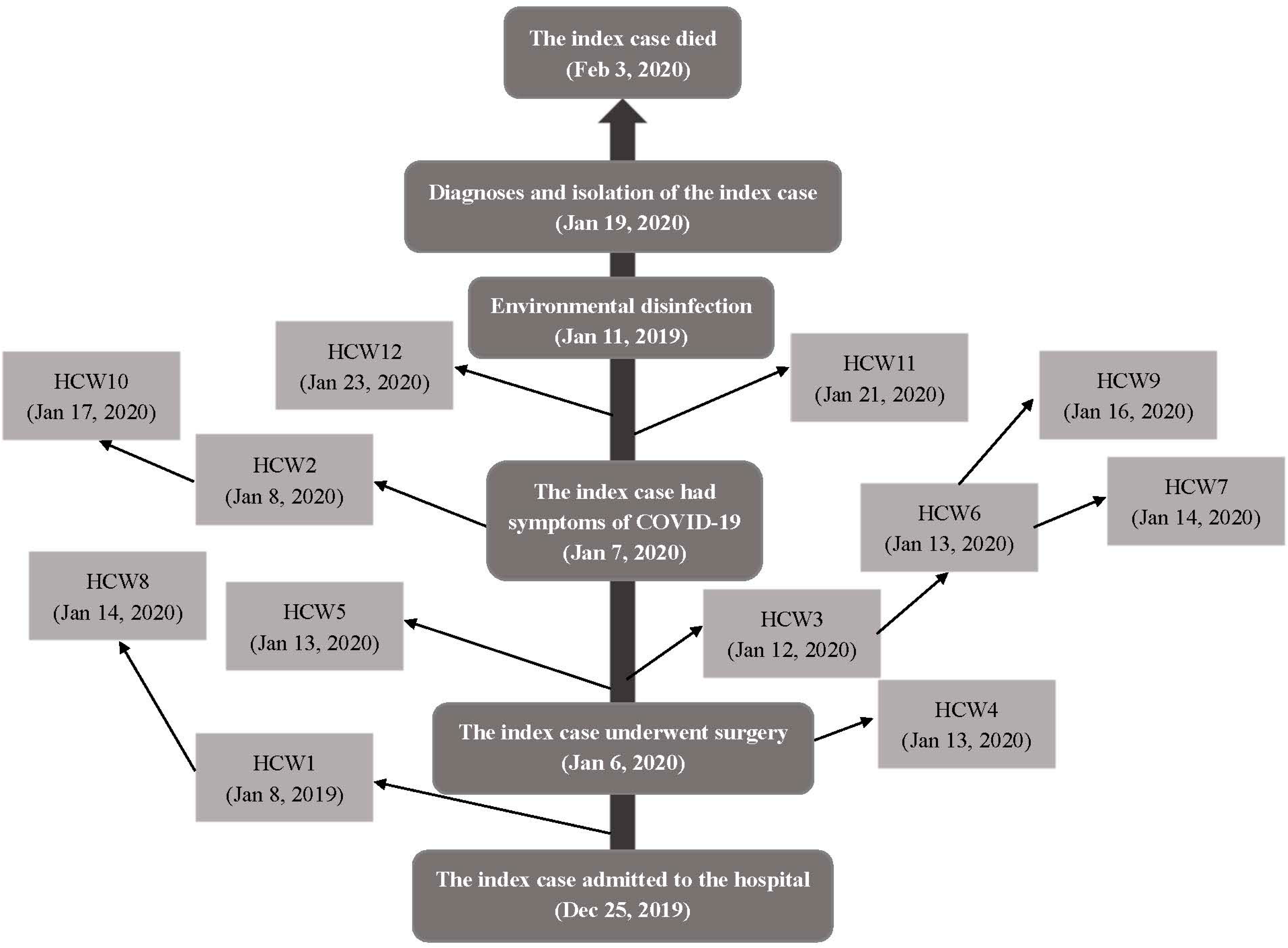
Overview of the transmission of COVID-19 from the index case to 12 healthcare workers. **Note:** HCWs, healthcare workers. The number (1∼12) for each HCWs was sorted according to the onset time (the date in parentheses) of their symptoms of COVID-19. Index case, the patient who was diagnosed with pituitary adenoma at first, and finally diagnosed with COVID-19, and this case was believed to be the source of infection among HCWs.

### Data Collection and Assessment

An online electronic questionnaire was sent to all 171 HCWs in the Department of Neurosurgery of Union Hospital of Wuhan, and 118 valid questionnaires were finally collected, including the questionnaires from 12 COVID-19 HCWs, and 106 uninfected HCWs. Baseline demographics (age, gender, height, weight, education level), lifestyle factors (physical activity, smoking status, and alcohol drinking status, diet), medical post, and chronic medical diseases were gathered. For all HCWs, their data on sleep quality were assessed by Pittsburgh Sleep Quality Index (PSQI),^10^ and for nurses, their feeling of working under pressure was further evaluated by The Nurse Stress Index (NSI).^11^ The contact status with the COVID-19 cases was also collected. A detailed description of these data as described in the Supplementary Materials. The follow-up duration for each HCWs was calculated as the number of days between Dec 25, 2019 (the hospital admission time of the index case) and the time when HCWs developed symptoms of COVID-19 or Feb 15, 2020.

### Statistical analysis

#### Risk Factors in The Susceptibility for COVID-19 in HCWs

Continuous variables were described as mean ± SD, or median and interquartile range (IQR), and values between COVID-19 HCWs and uninfected groups were compared using independent Student’s *t*-test or Mann-Whitney *U* test when data were normally or skewed distributed, respectively. Categorical variables were described using No. (%) and were compared using the χ2 test, or the Fisher’s exact test. Hazard ratios (HRs) and 95% confidence intervals (CIs) for the risk of COVID-19 were calculated by Cox proportional hazards regression models, with adjustment for age, gender and medical post (if necessary) in model 1, while HCWs’ exposure status to COVID-19 patient or HCWs were additionally adjusted in model 2. All the statistical hypothesis tests were two-sided with p-value < 0.05 as the level to reject the null hypothesis, and these analyses were performed with the SAS program (version 9.4; SAS Institute, Carry, NC)

### Individual-Based Model

Detailed procedures of the construction of IBM were described in the Supplementary Materials. In our model, a host can undergo a sequence of infection states, classified as susceptible (S), exposed (E)-infected, but not yet infectious, asymptomatic/mild symptomatic infectious (A), symptomatic infectious (I), or quarantined/isolated at home/recovered state (R) (see Figure S1). We assume that once an HCW becomes symptomatic, he/she will be quarantined or isolated at home as soon as possible, on average 1 day past COVID-like symptoms, depending on the heterogeneous level of symptomatic.

The key inputs in the model include 1) initial population makeup and infection status; 2) infectivity levels for asymptomatic/mild and symptomatic infections; 3) susceptibility levels of different hosts; 4) average duration of each state; 5) external source from infectious patients; 6) contact mixing patterns between hosts on daily basis (Table S2); The latter is simply defined as the number of daily contacts between HCWs. The model was implemented and simulated in Wolfram Mathematica 12.

## Results

### Presenting Characteristics

The mean age of COVID-19 HCWs was 36.6 (SD=7.4) years old, which was significantly higher than 106 uninfected HCWs (30.5 ± 5.3 years old) (Table 1). A high proportion of COVID-19 HCWs had a master’s degree or above, engaged in night shift-work (75.0% vs. 40.6%), felt they were working under pressure (66.7% vs. 32.1%), and had ever contacted the index case or COVID-19 HCWs (100.0% vs. 28.3%) than uninfected HCWs. The distributions of other demographical characteristics (sex, BMI), lifestyles (smoking status, alcohol drinking status, physical activity, and diet), the status of contacting the index case, and chronic medical disease were not significantly different between COVID-19 HCWs and uninfected HCWs.

**Table 1.** General characteristics of healthcare workers.

| Variables | All HCWs<br>(n=118) | Uninfected HCWs<br>(n=106) | COVID-19 HCWs<br>(n=12) | p value |
| --- | --- | --- | --- | --- |
| Age, years |  |  |  |  |
| Mean (SD) | 31.1 ± 5.8 | 30.5 ± 5.3 | 36.6 ± 7.4 | **0.006** |
| Range | 23~51 | 23~50 | 27~51 |  |
| Sex |  |  |  |  |
| Men | 43 (36.4) | 38 (35.9) | 5 (41.7) | 0.76 |
| Women | 75 (63.6) | 68 (64.1) | 7 (58.3) |  |
| BMI (kg/m<sup>2</sup>) | 22.1 ± 3.3 | 22.0 ± 3.3 | 22.4 ± 3.7 | 0.85 |
| Education level |  |  |  |  |
| College or university graduate | 92 (78.0) | 86 (81.1) | 6 (50.0) | 0.024 |
| Graduate or above | 26 (22.0) | 20 (18.9) | 6 (50.0) |  |
| Current smoking | 9 (0.08) | 9 (8.5) | 0 (0) | 0.63 |
| Current alcohol drinking | 10 (0.08) | 10 (9.4) | 0 (0) | 0.57 |
| Regular physical activity | 40 (33.9) | 34 (32.1) | 6 (50.0) | 0.33 |
| Regular diet | 53 (44.9) | 47 (44.3) | 6 (50.0) | 0.77 |
| Number of daily diets | 2.7 ± 0.5 | 2.7 ± 0.5 | 2.8 ± 0.4 | 0.71 |
| Medical post |  |  |  | 0.52 |
| Nurse | 88 (74.6) | 80 (75.5) | 8 (66.7) | 0.019 |
| General nurses | 31 (35.2) | 29 (36.3) | 2 (25.0) |  |
| Nurse practitioners | 42 (47.7) | 41 (51.2) | 1 (12.5) |  |
| Nurse-in-charge | 15 (17.1) | 10 (12.5) | 5 (62.5) |  |
| Doctor | 30 (25.4) | 26 (24.5) | 4 (33.3) |  |
| Night shift-work |  |  |  |  |
| No | 66 (55.9) | 63 (59.4) | 3 (25.0) | 0.023 |
| Yes | 52 (44.1) | 43 (40.6) | 9 (75.0) |  |
| Working under pressure |  |  |  |  |
| No | 76 (64.4) | 72 (67.9) | 4 (33.3) | 0.022 |
| Yes | 42 (35.6) | 34 (32.1) | 8 (66.7) |  |
| Contact the index case <sup>a</sup> |  |  |  |  |
| No | 22 (18.6) | 17 (16.0) | 5 (41.7) | 0.077 |
| Yes | 96 (81.4) | 89 (84.0) | 7 (58.3) |  |
| Contact mode |  |  |  |  |
| Air | 35 (36.5) | 32 (36.0) | 3 (42.9) | 0.90 |
| Direct contact | 49 (51.0) | 46 (51.7) | 3 (42.9) |  |
| Both | 12 (12.5) | 11 (12.4) | 1 (14.3) |  |
| Contact frequency (No./day) | 5.0 (2.0, 6.0) | 5.0 (2.0, 6.0) | 3.0 (1.0, 6.0) | 0.95 |
| Average contact duration (min/time) | 4.0 (2.0, 6.0) | 4.0 (2.0, 6.0) | 4.0 (2.0, 6.0) | 0.54 |
| Longest contact duration (min) | 10.0 (5.0, 25.0) | 10.0 (5.0, 20.0) | 10.0 (5.0, 30.0) | 0.69 |
| Contact the index case or infected HCWs <sup>b</sup> |  |  |  |  |
| No | 76 (68.6) | 76 (71.7) | 0 (0) | <0.001 |
| Yes | 42 (31.4) | 30 (28.3) | 12 (100.0) |  |
| Chronic medical disease |  |  |  |  |
| Pulmonary disease | 9 (7.6) | 8 (7.6) | 1 (8.3) | 0.49 |
| Non-pulmonary disease | 6 (5.1) | 6 (5.7) | 0 (0) |  |

**Note:** Continuous variables were expressed as mean ± SD or median (IQR), and compared by Student's *t*-test or the Mann-Whitney *U* test, respectively. Categorical variables were expressed as No. (%) and compared by the $\chi^2$ test or Fisher's exact test.
<sup>a</sup> The patient who was initially hospitalized with pituitary adenoma, and finally diagnosed with COVID-19.
<sup>b</sup> The index case and COVID-19 HCWs in the same department.

### Distribution of The Pittsburgh Sleep Quality Index and The Nurse Stress Index

Given the findings that a higher proportion of COVID-19 HCWs worked the night shift and felt they were working under pressure than uninfected HCWs, we further evaluated their sleep quality and working pressure by PSQI and NSI. The analyses showed that COVID-19 HCWs had a significantly higher score of PSQI than uninfected HCWs (p value<0.001, Figure 2A). Specifically, for the 7 factors PSQI test, COVID-19 HCWs had significantly high scores for 5 factors (sleep quality, sleep time, sleep efficiency, sleep disorder, and daytime dysfunction), while the other 2 factors (sleep duration, and use of the hypnotic drug) were not significantly different. For the NSI conducted among nurses, the scores of its 5 subscales (nursing profession and work, workload and time allocation, working environment and resources, patient care, management and interpersonal relations) were all significantly higher in infected than uninfected nurses (Figure 2B).

**Figure 2.**
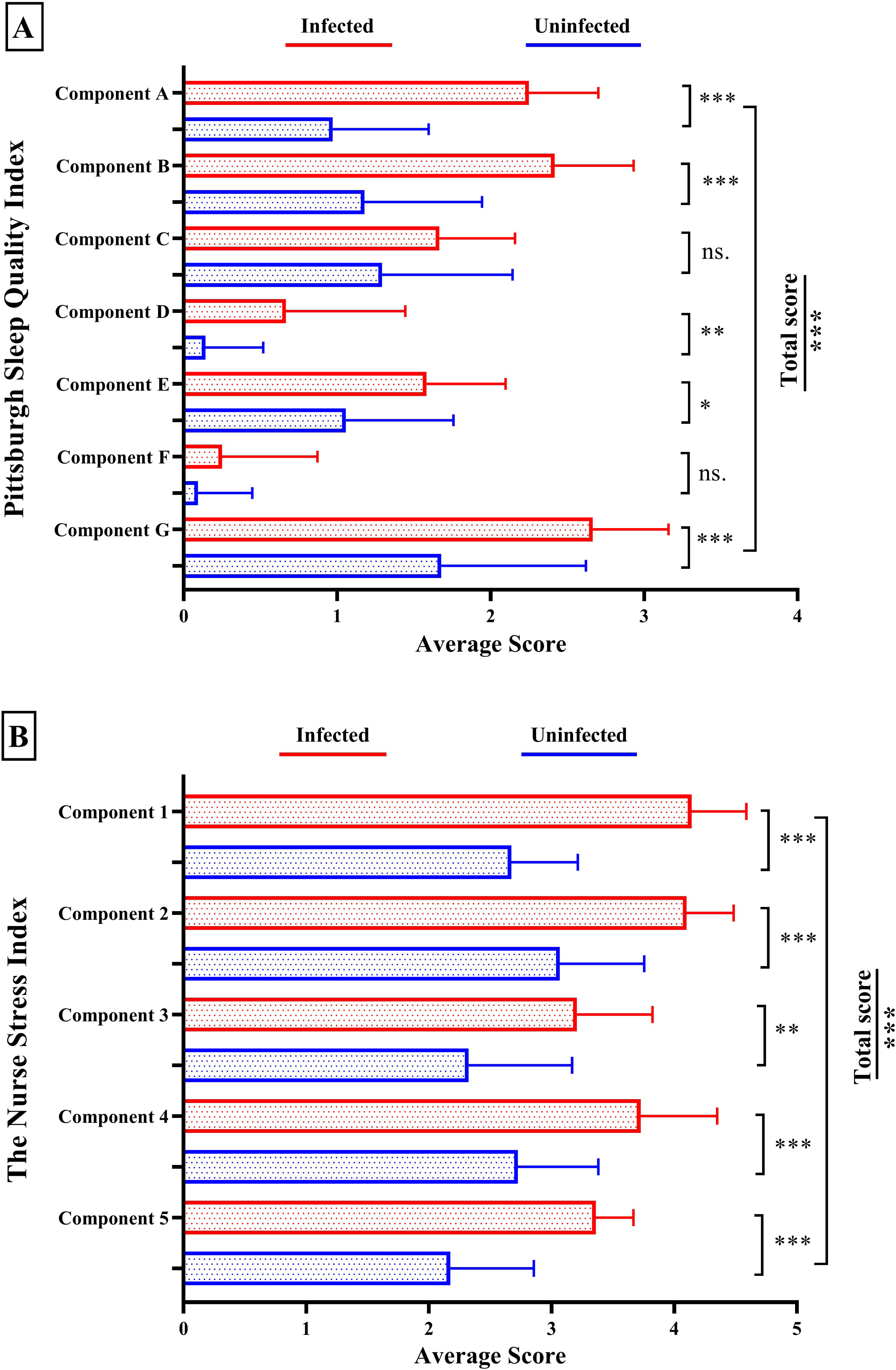
The difference in the distribution of the Pittsburgh Sleep Quality Index and The Nurse Stress Index between COVID-19 cases and uninfected healthcare workers. **Note:** Figure A: Component A, sleep quality; Component B, sleep time; Component C, sleep duration; Component D, sleep efficiency; Component E, sleep disorder; Component F, hypnotic drug; Component G, daytime dysfunction; Total score, summary values of the 7 factors from A to G. Figure B: Component 1, the stress of Nursing profession and work; Component 2, the stress of Workload; Component 3, the stress of Working environment and resources; Component 4, the stress of Patient care; Component 5, the stress of Management and interpersonal relations. The total score, summary values of the 5 subscales from 1 to 5. The straight bar is the mean score of each component in scale and the whisker line is the standard error. HCWs, healthcare workers. ns., not significant. p-value * <0.05; ** <0.01; *** <0.001.

### Associations of the Pittsburgh Sleep Quality Index and The Nurse Stress Index with The Risk of COVID-19

We further investigated the associations of sleep quality and working pressure with the risk of COVID-19. The total score of PSQI and NSI were positively associated with the risk of COVID-19 (Table 2), no matter if the contact status to the index case or infected HCWs were adjusted (model 2) or not (model 1). For PSQI, high scores on the factors of sleep quality and sleep time were associated with high risks of COVID-19, and for NSI, high scores on the subscales of the nursing profession and work, and management and interpersonal relations were associated with high COVID-19 risk.

**Table 2.**
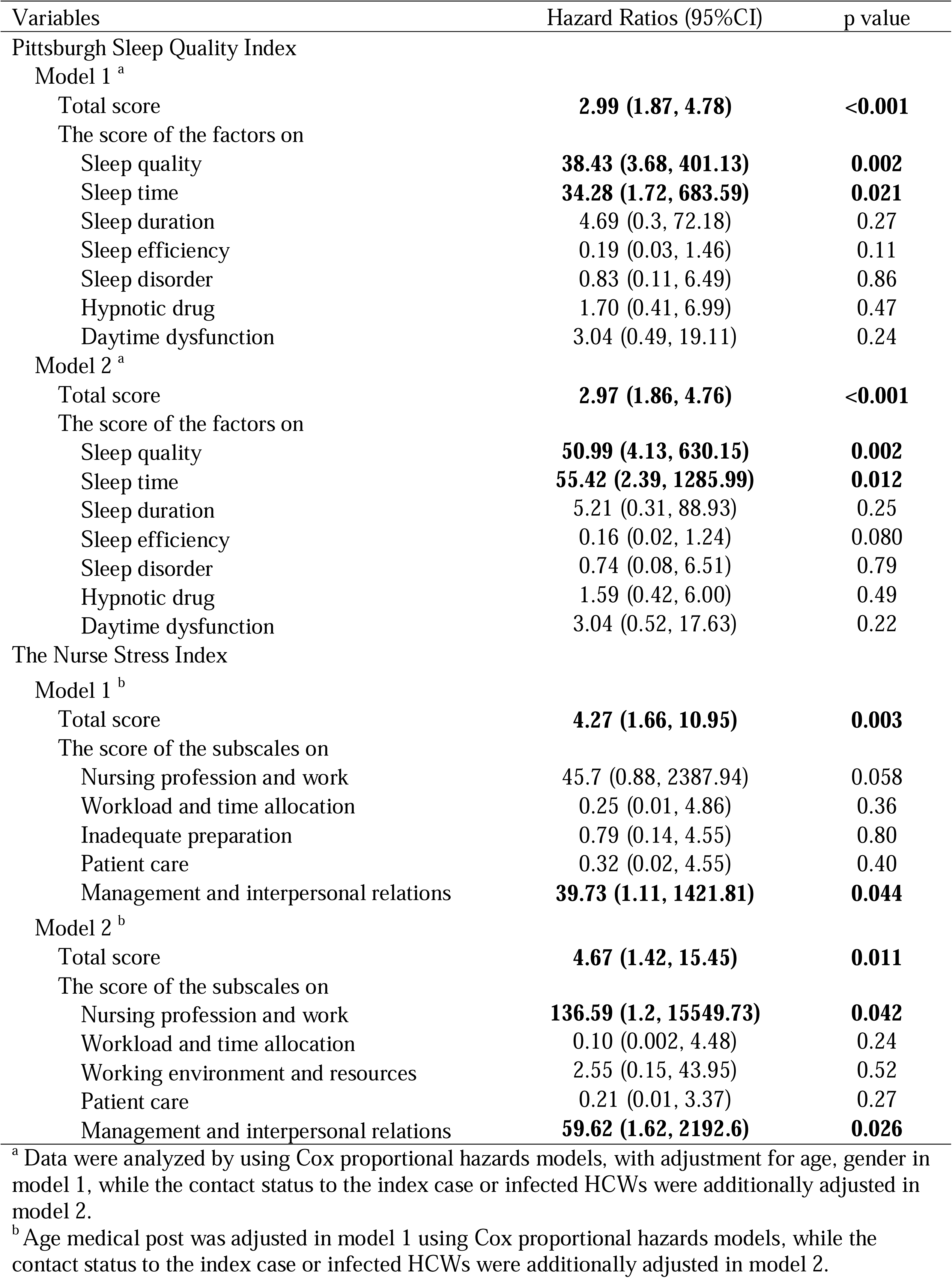
Associations of the Pittsburgh Sleep Quality Index and The Nurse Stress Index with the risk of COVID-19.

### Model Simulations

To explore the transmission pattern of the COVID-19, we simulated the IBM transmission dynamic of COVID-19 like a pathogen, in a host pool of 118 HCWs over 120 days. We initialized our system with a single infected host, in a susceptible pool. The mixing pattern was based on the prescribed number of pairs and triple contacts among HCW and their contacts with the prescribed patient pool.

We considered two cases, e.g., (i) no patient contacts; (ii) external patient-contact source for HCW. Other parameter values are outlined in Table S2.

In case (I), Figure S2 showed that the epidemic peak was reached after 80-100 days, the highest incidence happened after 75 and 95 days with 3 newly infected HCWs on those two days, and in total there would be about 64 HCWs being infected after 120 days. *R*_0_ was estimated to be 1.18 [1.15, 1.21].

Case (II) added external patient-source (Figure 3). Here epidemic peak came earlier (62-80 days), and reached a higher incidence level (4 additional HCWs on certain days). The total number of quarantined/recovered/isolated HCWs after 120 days has increased to 80. The resulting *R*_0_ from model simulation was estimated to be 1.27 [1.06, 1.61]. Our simulation fitted the first 30 days’ data well (12 infected HCWs among 118 HCWs from Dec 25, 2019 to Jan 23, 2020 in Table S1). *R*_0_ was estimated to be 1.03 from this limited outbreak data.

**Figure 3.**
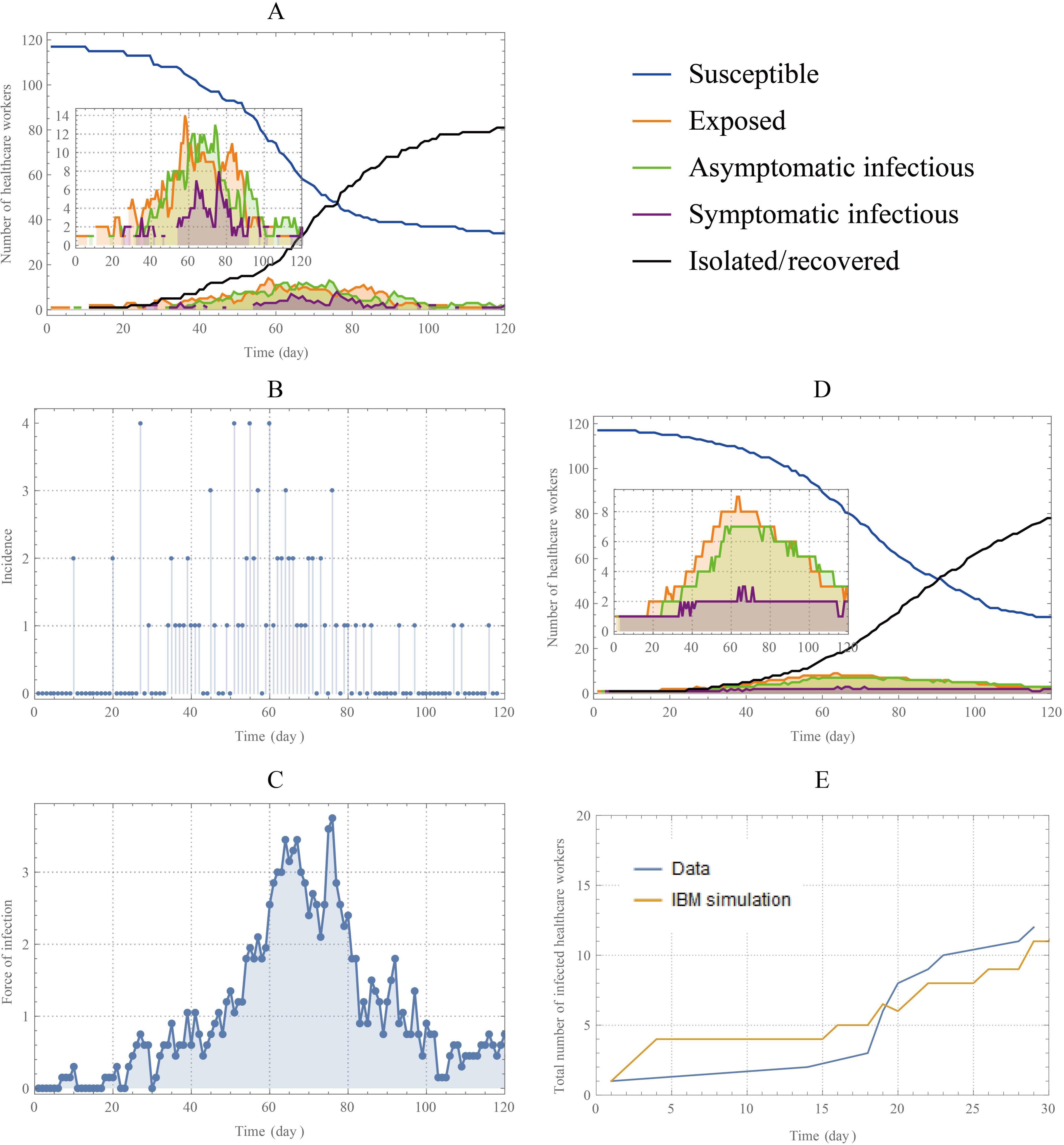
Typical IBM history simulations for a community of 118 hosts with external source. **Note:** Panel (A) shows the typical history for five different classes (susceptible, exposed, asymptomatic/mild infectious, symptomatic infectious and quarantined/isolated at home/recovered) by IBM simulations for 120 days with external source (contact with infectious patient), based on the parameter values and initial conditions listed in Table S2; Panel (B) gives us the force of infections; Panel (c) shows us the incidence (the number of newly infected cases of COVID-19); Panel (d) is the median solutions of 200 IBM simulations. Panel (E) is the comparison between the data and the median solutions of 200 IBM simulations.

### Preventive Measures and Control Interventions

Host susceptibility level (and the resulting community transmission rate) could be reduced by additional preventive means, e.g. personal protective equipment (PPE)and washing hands. PPE shortages have been described in the most affected facilities. Hence, in a non-typicalCOVID-19 ward, there is high probability HCW staff has insufficient protective gear (e.g. masks), or they may lack awareness.

Additional risk factors and possible preventive steps include reasonable assignment of workload or immediate stress relief for HCWs we mentioned above.

Especially, health-care workers are anxious about passing the infection to their families at this moment. Governments or hospitals should try to arrange appropriate accommodations for those HCWs who had this kind of concerns. Quantifying the effect of such preventive means on susceptibility is a challenging task. However, by reducing the susceptibility of HCWs by a 1.40 factor during simulation, we observed the epidemic peak occurring on day 80 with a combined infected pool of 40 over 120 days and the highest incidence happened with 2 newly infected hosts (Figure 3A).

Further, hospitals should also change some non-urgent meetings to remote meetings. By combined control strategies (reducing the average contact rate per HCW by a 1.35 factor and reducing the susceptibility of HCWs by a 1.40 factor), we were able to reduce the total infected pool to 10, and thus avoided a big outbreak of the basic case (Figure 3B).

## Discussion

As the pandemic (COVID-19) accelerates, millions of people are recommended to work from home (social distancing) to minimize the transmission of COVID-19, health-care workers are doing the exact opposite -going to hospitals, clinics, and putting themselves at high risk from COVID-2019 ^5^.

The present study was conducted in a department of neurosurgery, which lacks the established practices of infection control, such as early detection and isolation, contact tracing and the use of personal protective equipment when compared with the department of respiratory medicine and infectious disease. Although the incident COVID-19 rate (8.19%) in this study was much lower than the rates for HCWs with SARS (19%) in Beijing, and MERS in Saudi Arabia (13.4%), respectively,^12^ it may be much reduced when the risk factors were investigated and controlled.

In this study, 7 of 12 COVID-19 HCWs were not directly contacted the index case (Figure 1), suggesting the human-to-human transmission between HCWs, which leads to the serious nosocomial infection. Although the main reason for this early infection among HCWs in hospitals was lack of sense of protection, the incident HCWs infection continue to occur after they had worn protective equipment,^4^ due to the contribution of other risk factors on HCWs infection.

The data in the present study suggested that a high proportion of COVID-19 HCWs worked the night shift. Furthermore, the PSQI showed a higher total score, sleep quality score and sleep time score among infected than uninfected HCWs, and these scores were positively associated with the risk of COVID-19. Although the underlying mechanism for these associations had not been explored, proper sleep is at the first line of defense against infections that had been reported^13^. Besides, findings in this study showed that a high proportion of COVID-19 nurses felt they were working under pressure, especially the pressure of dealing with a pneumonia of unknown etiology, such as COVID-19. We further analyzed the pressure source by NSI, and these scores were positively associated with COVID-19 risk among nurses when the contact status with infected cases was adjusted. One possible reason is the prevalent of oxidative stress among nurses with higher job stress,^14^ which can damage the immune function,^15^ and further lead to the increased susceptibility to COVID-19. However, due to the retrospective design of the present study, the data of the individual’s immunity before their infection was not collected, and the hypothesis needs to be further validated.

Until now, no study has evaluated the risk factors that may play roles in the susceptibility of COVID-19 among HCWs before the outbreak of COVID-19. In this study, we collected the potential risk factors before the measures of infection control were widely conducted, which ensure the data was under the natural transmission of COVID-19. Besides, we used a novel IBM to simulate the dynamics transmission of COVID-19 spread through person-to-person contact among HCWs, which accounted for the heterogeneity of disease progression, contact patterns, and transmission rates, when compared with the regular SEIR models. Our IBM simulation fitted the first 30 days’ data well (12 infected HCWs among 118 HCWs from Jan 8, 2019, to Jan 23, 2020) and estimated that the epidemic peak was reached after 62-80 days with a high incidence level (4 additional HCWs on certain days). The total number of infected HCWs after 120 days could increase to 80 if no control implications. The resulting R_0_ was estimated to be 1.27 (range [1.06, 1.61]), from model simulations, and *R*_0_= 1.03, from data analysis (though the outbreak data is very limited). However, if we implement the control strategies mentioned above (e.g., enough personal protective equipment even in a non-typical COVID-19 ward, decreasing contact among HCWs, and providing reasonable assignment of workload or immediate stress relief for HCWs), we were able to avoid an outbreak among HCWs. We should remember that the safety of HCWs must be ensured and health-care workers are most valuable resources to fight for COVID-19.

There are limitations and future studies. First, there was the possibility of unmeasured residual confounding effects of contact status with infected cases, although we had adjusted for some primary confounders. Second, the insufficient sample size may influence the statistical power. Further large prospective studies are needed to validate our findings. Third, we ignored dynamic interactions between patients and HCWs in our IBM, but simply considered infectious patients as external sources. A more comprehensive model including patients, visitors, nurses, doctors, staff, and even family members should be studied in the future since the hospital is not a closed system. Fourth, our model has many uncertain parameters (susceptibility, infectivity levels, disease progress, and detailed contact patterns) that could be adjusted to available data for any given local community and can be further used to explore the efficacy of different control strategies for specific local communities and single entities (e.g., schools, working places, hospitals).

## Conclusion

The data before the outbreak of COVID-19 showed not enough personal protective equipment, poor sleep quality and high working pressure were positively associated with high risks of COVID-19. An individual-based model of COVID-19 transmission in a hospital setting is suitable for simulating different outbreak patterns. These results provided epidemiological evidence on the susceptibility of HCWs to COVID-19 and a method for assessing the transmission model of COVID-19 among HCWs.

## Data Availability

At presentϊ the data is confidential.

## Declaration of Interest

None exist.

## Funding/Support

This work was supported by the Fundamental Research Funds for the Central Universities (grant 2020kfyXGYJ010 to Dr. X. Jiang).

## Role of the Funding Source

The funders had no role in the design and conduct of the study; collection, management, analysis, and interpretation of the data; preparation, review, or approval of the manuscript; and decision to submit the manuscript for publication.

## Author Contributions

Y.B., W.X, and Q.H contributed equally and share first authorship. X.J. and H.Z. contributed equally to this article. Y.B., W.X, Q.H, and H.W. analyzed data and wrote the paper. Q. H, D.G., M.M, and M.H did model development and simulations, D.G., M.M, M.H, S.X. and A.M. made critical revision of the manuscript. X.J. and H.Z conducted research. Y.B., W.X, Q.H, H.W., and X.J. designed research. R.L., H.S., F.F., and P.F. collected and provided essential materials.

## Acknowledgment

The authors would like to appreciate all healthcare workers in this study. X.J. and X.W. had full access to all the data in the study and takes responsibility for the integrity of the data and the accuracy of the data analysis. All authors had commented on the report drafts and approved the final submitted version. All authors were free from potential conflicts of interest.

## Notes

### Competing Interest Statement

The authors have declared no competing interest.

